# Chiari malformation does not obstruct cerebrospinal fluid flow during coughs

**DOI:** 10.1101/2025.05.01.25326708

**Authors:** Robert A. Lloyd, Adam D. Martinac, Zhaoyan Liu, Joel A. Berliner, Marcus A. Stoodley, Lynne E. Bilston

## Abstract

Chiari malformation is characterised by the herniation of the cerebellar tonsils through the foramen magnum. It is commonly assumed that the tonsils obstruct normal cerebrospinal fluid (CSF) flow, resulting in cough-associated headaches. However, the mechanisms that cause these headaches are unknown, leading to varied and often unsatisfactory treatment outcomes for patients with Chiari. In this study real-time phase contrast MRI (PC-MRI) was used to determine whether the tonsils restrict CSF flow during coughs, and shape models of the posterior fossa were used to identify morphological differences between patients with and without cough headache. Twenty Chiari patients with cough headache, seven without, and twelve age and sex matched controls underwent MR imaging of the head and neck. 3D models of the hindbrain, CSF and vertebrobasilar arteries were constructed from T1-weighted MRI. PC-MRI was collected at the foramen magnum and mid-C3 to measure CSF flow, plus arterial and venous flow. The shape models showed that the CSF space anterior to the pons was narrower in Chiari patients than in controls. Additionally, the CSF space around the vertebrobasilar arteries was more restricted in patients with cough headache than those without. The flow of CSF across the foramen magnum was not restricted, and CSF velocities were ∼2 times greater in Chiari patients than in controls. These findings suggest that overcrowding of the posterior fossa creates high velocity CSF flow across the pons, medulla, and vertebrobasilar arteries. This high velocity flow through the restricted CSF space may stress the cranial vasculature and contribute to headache.

**Highlights:**

- The Chiari patients CSF flow was not blocked during coughs
- Patient cough induced CSF velocities were 2 times greater than controls
- Shape models showed reduced CSF space anterior to the pons in patients with Chiari
- Cough headache patients had greater CSF restrictions around the cranial arteries

## Introduction

Chiari malformation Type I (hereafter Chiari) is a congenital disorder defined by overcrowding of the hindbrain and descent of the cerebellar tonsils through the foramen magnum into the cervical spinal canal (Figure 1)(Milhorat et al., 1999). Most people living with Chiari experience debilitating headaches, which are typically (but not universally) suboccipital and triggered by coughing, straining, or physical exertion (Alperin et al., 2015). How Chiari leads to headache is unknown (Hersh et al., 2019), and traditional imaging markers such as the depth of tonsillar herniation are not related to a persons’s symptoms (Meadows et al., 2000; Thakar et al., 2021).

**Figure 1.**
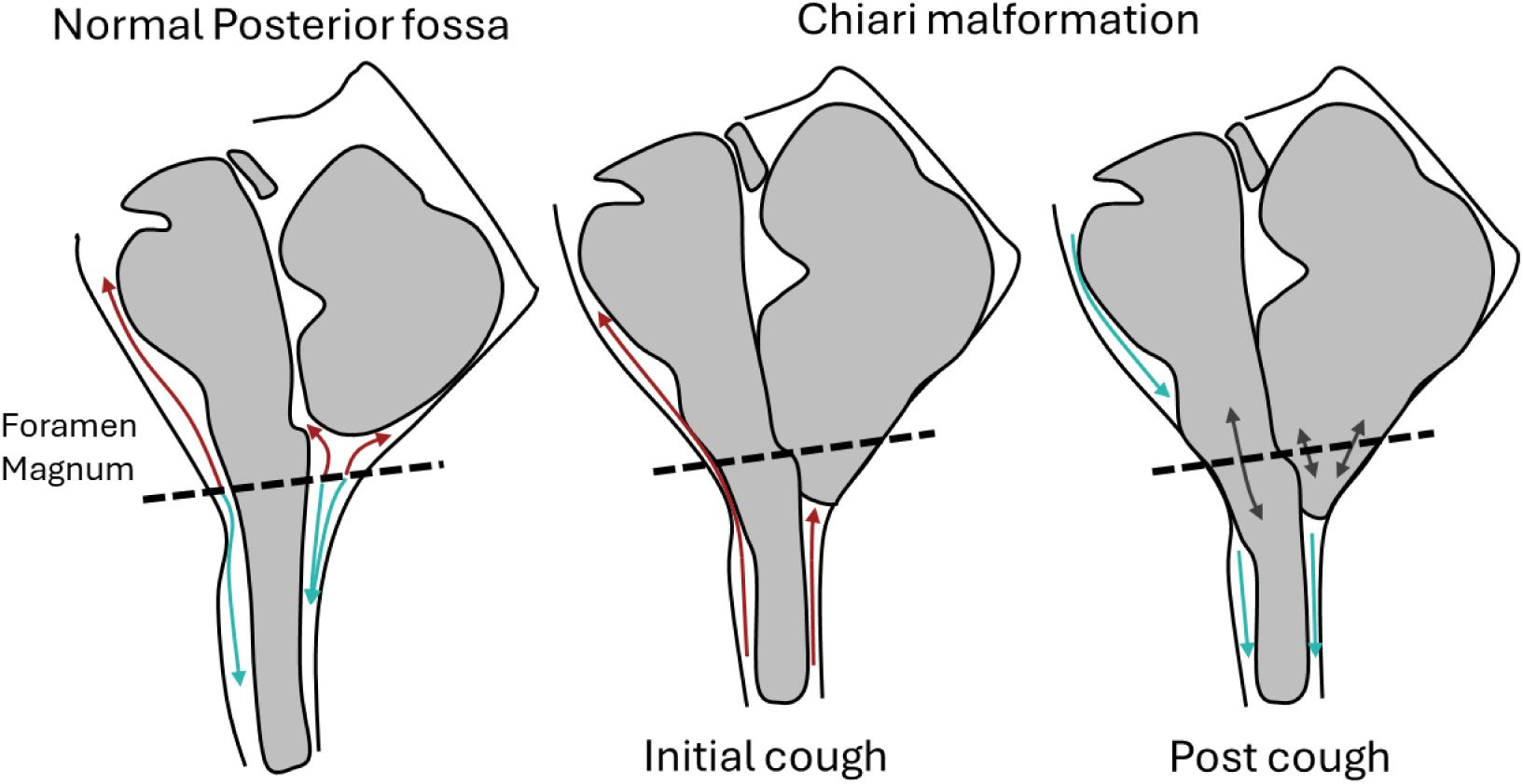
Schematic of the effects of coughs on CSF flow. A) Normally the posterior cranial fossa is not obstructed by the cerebellum, and CSF is free to flow cranially during a cough (red arrows) and return caudally (blue arrows) post cough. In patients with Chiari, it is hypothesised that CSF flows freely into the cranium during a cough (B), but the cerebellar tonsils obstruct the caudal return of fluid post cough (C). This is thought to stretch nervous tissues (black arrows) triggering headaches.

In healthy individuals, a cough rapidly increases intrathoracic and intra-abdominal pressures (Lloyd et al., 2020). This leads to an influx of venous blood into the spinal canal, and increases the caudal spinal pressures (Usubiaga et al., 1967), displacing CSF cranially (Figure 1A) (Du Boulay et al., 1972; Lloyd et al., 2020). After the cough, intrathoracic and abdominal pressures decrease, and CSF returns caudally (Figure 1A). As Chiari headaches are commonly associated with coughing, it has often been assumed that the herniated cerebellum alters the pattern of CSF flow during a cough, and this triggers the headaches (Bezuidenhout et al., 2019). The suggested mechanism is that overcrowding of the posterior fossa forms a partial obstruction to CSF flow (Williams, 1980). During the early phase of the cough the CSF flows into the cranium (Figure 1B), but as the cough-induced transient pressure increase ends, the hindbrain is displaced caudally and restricts the outflow of CSF (Figure 1C), increasing CSF pressure above the foramen magnum, and stretching neural tissues which triggers headache (Dawes et al., 2019; Leung et al., 2016). This hypothesis underpins the mainstay of clinical management of Chiari: posterior fossa decompression, where the space around the hindbrain is surgically enlarged and CSF flow is supposedly normalised. However, posterior fossa decompression is unsuccessful in resolving headaches in 26-46% of cases (Parker et al., 2013; Wang et al., 2023; Zhou et al., 2024), which calls into question the current mechanistic understanding of Chiari headaches.

Recently, Bhadelia et al. (2024) used real-time pencil-beam phase contrast MRI (PC-MRI) to determine whether CSF flow during and after coughing was different between healthy controls and Chiari patients with or without cough-associated headaches. In Chiari patients with cough headaches, they found CSF pulsation was reduced after six coughs of maximal effort. This was taken as evidence that the cerebellum obstructs CSF flow, in support of Williams’ (1980) original hypothesis. However, repeated forceful coughs have been shown to trigger a drop in arterial pressure, (Sharpey-Schafer, 1953) and elevate the heart rate for 10 – 20 seconds after coughing (Wei and Harris, 1982; Wei et al., 1983). This coupled cardiovascular response would also affect the cranial CSF pulses, which may explain the variation in the post-cough changes measured 5-10 seconds after coughing by Bhadelia et al. (2024). To date the effects of coughing in Chiari have only been assessed by pencil beam PC-MRI (Bezuidenhout et al., 2018; Bhadelia et al., 2024; Bhadelia et al., 2016), which provides a one-dimensional measure of flow, and assumes there to be negligible spinal cord movement. It is unknown how the morphology of the posterior fossa influences the patterns of CSF flow during cough.

The aim of this study was to use anatomical MRI to characterise the morphology of the posterior fossa in people with Chiari and to use real-time PC-MRI to measure the effect of single coughs on CSF flow. We hypothesise that the reduced anterior CSF space in patients with Chiari headache would have the fastest cranial CSF velocities, but the lowest CSF velocities during the caudal return after the cough.

## Methods

### Ethical Approval

The South Eastern Sydney Local Health District Human Research Ethics Committee approved all experimental protocols, and they were conducted according to the Declaration of Helsinki (2013) except for registration in a public database. All participants gave written informed consent.

### Subjects

Forty-five participants with no contraindications for MRI underwent imaging of their head and neck. Six were excluded from analysis due to poor image quality. The cohort consisted of 27 people with symptomatic Chiari malformation type I (Age: 36±9 years, BMI: 27±6 kg.m^-2^, 1 male), and 12 age and sex matched healthy controls (Age: 31±8 years, BMI: 22±2 kg.m^-2^, 1 male). All 27 Chiari participants reported experiencing suboccipital headaches, 20 of which experienced headaches that were triggered or worsened by coughing, straining, or exercise.

### MRI Scans

MRI data were collected using a Philips Ingenia 3CX (Philips Healthcare, Best, The Netherlands), with a 16-channel head and neck coil. Participants were imaged supine, with the Frankfort plane vertical. For anatomical scans participants were instructed to keep still and breathe quietly. During the real-time phase contrast (PC) MRI, participants were instructed to cough gently after the expiratory phase of each breath. Respiratory motion was recorded concurrently with a respiratory sensor positioned just below the xiphoid process.

T1 weighted scans were acquired to extract the morphology of the posterior cranial fossa, hindbrain, and the cerebral arteries (vertebral, and basilar arteries). Scan parameters included TR/TE = 8.2/3.74 ms, matrix = 256×256, 180 sagittal slices, voxel size = 1×1×1 mm^3^.

Real-time PC-MRI was used to collect CSF and blood flow data during respiratory manoeuvres. The real-time method was not gated and acquired phase images at the fastest rate achievable with the given scan parameters. The timing of images was synchronised with respiratory manoeuvres using the respiratory band signal. Imaging planes were positioned at mid-C3, or near the foramen magnum. In controls scans were collected at the foramen magnum. In Chiari patients the plane was placed 5mm above the tip of the tonsils, avoiding the CSF space where the vertebral arteries crossed the anterior space. Scanning parameters included: flip angle = 20°, matrix=96×81, FOV=192×192 mm, TR/TE = 13/7 ms, Slice thickness: at C3 = 7.5 mm, at the foramen magnum = 5 mm. The encoding velocities were set to 10 – 45 cm s^-1^ for CSF and 55 – 105 cm s^-1^ for arterial and internal jugular flow, acquiring 200 phases at 70 and 140 ms intervals. Scans were repeated with a higher encoding velocity when aliasing artefacts were present. Small aliasing errors that were present were corrected with a 3D phase-unwrapping algorithm (Abdul-Rahman et al., 2005; Bydder, 2019).

### Image segmentation

Regions of interest (ROIs) were manually segmented using 3D slicer (https://www.slicer.org, (Fedorov et al., 2012)) by a single researcher (RL). The CSF space and nervous tissues from mid-C5 in the spinal canal to the tentorium of the posterior cranial fossa were segmented on the anatomical T1-weighted images (Figure 2A&B). Additionally, the vertebral and basilar arteries within the posterior fossa were also segmented. Separate ROIs were extracted for the CSF spaces from the PC-MRI (Figure 2C&D). At C3, ROIs for the vertebral and internal carotid arteries, and internal jugular veins (IJV) were segmented. These ROIs were used to iteratively train a segmentation model using the nnU-Net framework (Isensee et al., 2021). Segmentation accuracy of the final model was tested against 20% of the labels that were withheld from training. The results are provided in the Supplementary material, and the trained models are available at [*Repository URL to be made before peer review*], which can be used with 3D slicer by other researchers.

**Figure 2.**
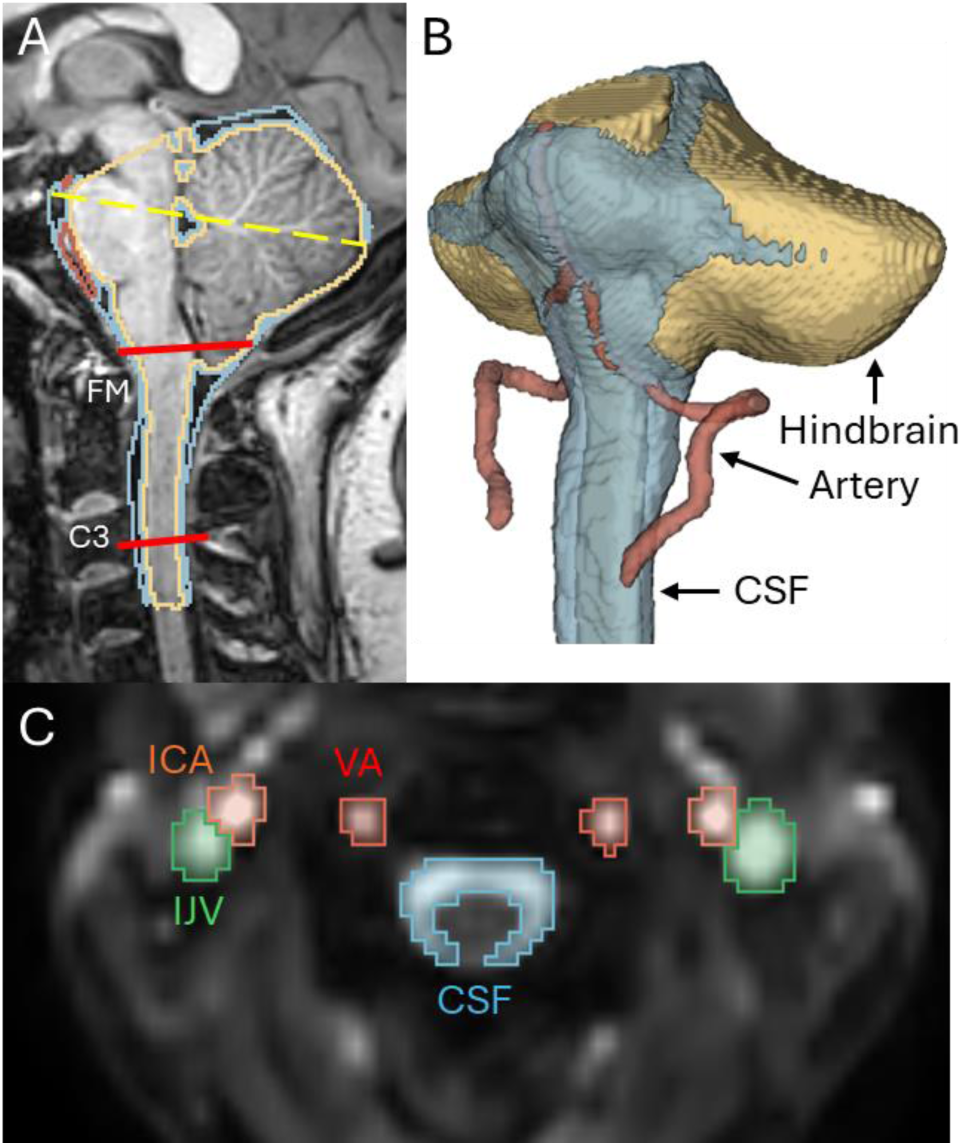
Example T1-weighted and PC-MRI scans showing the segmented ROIs. A) T1-weighted mid-sagittal slice. The CSF space is outlined in blue, the hindbrain in yellow and the arteries in red. The solid red lines indicate the imaging planes at the foramen magnum (FM), and mid-C3. The yellow dashed line between the clivus and internal occipital protuberance was used to select the superior CSF space for Procrustes analysis. B) 3D view of the segmented ROIs used for morphological analysis, with the same colour coding as A. C) shows the ROIs segmented from the PC-MRI at mid-C3. The ROIs are colour coded as follows, blue = CSF, red = vertebral arteries, orange = internal carotid arteries, and green = internal jugular veins.

### Morphological analysis

To compare differences in the posterior cranial morphology, the CSF, tissue, and vascular volumes were resampled as point-clouds. The CSF and hindbrain were resampled using ShapeWorks (V6.1, (Cates et al., 2017; SCI Institute, 2021)). The branches of the vertebrobasilar arteries were skeletonised and approximated by 11 ellipsoids fitted perpendicular to the vessel lumen. Procrustes analysis was used to align the superior cranial fossa of each subject (above the line connecting the clivus to the internal occipital protuberance; Figure 2A) to the median model. This removed variation in the position, rotation, and overall size between subjects. The same transform was applied to whole CSF, hindbrain, and vascular volumes to align these ROIs. The branches of the vertebral arteries are described to enter the CSF space laterally via the foramen magnum and converge at the midline to form the basilar artery. However, within the normal population, the curvature of the vessels and where they converge relative to the medulla oblongata vary. Differences in the left-right bias of the vertebrobasilar arteries may influence where the CSF is narrowest in patients with Chiari and should be accounted for. To prevent lateral curvature of the arteries being removed from the shape models by averaging, the participants’ data were mirrored in the sagittal plane, so all vessels were left-side biased. Differences in the structure of the CSF space were characterised by the normal distance between the vertices of the hindbrain, blood vessels and posterior cranial fossa.

### Flow analysis

CSF and blood flow rates (in mL.s^-1^) were calculated by numerically integrating the measured velocities over the area of each ROI. The velocities reported in this study are the average velocity of each ROI (or the flow rates/ROI area). The signal from the respiratory band was used to identify the timing of the flow with respect to the cough. For each scan, the time-series mean was calculated and peak CSF flow rates that were ±2 S.D. away from the time-series mean during coughs were analysed. The peak cranial and caudal CSF flow and velocity were recorded before, during, and after the coughs. The flow through the left and right blood vessels was summed to calculate total flow. Blood flow is reported as the peak systolic flow rate. During coughs, blood flow was reported as the peak flow rate at the end of the expiratory phase of the cough.

### Statistical analysis

Partial least squares (PLS) regression is a data reduction technique that calculates latent variables that characterise the largest source of covariance between the independent, and dependent variables. For a shape model, the latent variables (shape modes) characterise the direction in which a shape deviates from the average model, and the shape scores give the magnitude of the change. To characterise the variation in the structure of the CSF space, PLS discriminant analysis (DA) was used to identify the shapes associated with each participant group. To characterise 95% of the variance in the between subject groupings, ten latent variables were used to build the PLS-DA model. Differences in the shape modes between groups were assessed with the general linear model (GLM), and group-wise differences were tested with a post-hoc Bonferroni correction.

A linear mixed model (LMM) was used to compare whether coughs altered the measured CSF and blood flows or velocities, and if this was different between subject groups. The model included fixed effects for the subject groups, the respiratory phase (before, during, or after a cough), and their interaction. The LMM included a random intercept for each participant to account for within-subject similarity between repeated measurements taken and different time points. The maximum likelihood method was used during fitting to deal with unbalanced/missing data. Pairwise comparisons were performed with a Bonferroni correction, considering P < 0.05 to be statistically significant.

## Results

### Differences in CSF spaces

Ten-fold cross-validation found that ten latent variables were able to capture 68% of the shape variations within our cohort and accounted for 96% of the variation between Chiari patients with and without cough headache and healthy controls. This shape model encoded changes in the shape and volume of the CSF space of the posterior cranial fossa, the hindbrain, and the vertebrobasilar arteries. Of the ten shape modes used to build the model, only the first four were significantly different between subject groups (GLM; Mode 1 – p<0.0001, Mode 2 – p=0.0054, Mode 3 – p = 0.018, Mode 4 – p=0.004). Shape mode 1 described the largest difference in the position of the hindbrain. A lower score corresponded to a more herniated cerebellum, an anteriorly positioned brain stem, and a reduced anterior CSF space (Figure 3). Both patients with (CAH) and without (CHI) headache, had lower Mode 1 scores than healthy controls (GLM; Mode 1 CAH d = -3.01, p < 0.0001, no CAH d = -2.51, p < 0.0001). Shape modes 2 – 4 characterised smaller changes in shape of the herniated tonsils and the volume of CSF in the posterior CSF space inferior to the foramen magnum. These modes also captured a rotation in the sagittal plane that made either the pons or medulla closer to the boundary of the cranium (Figure 4: Sagittal view).

**Figure 3.**
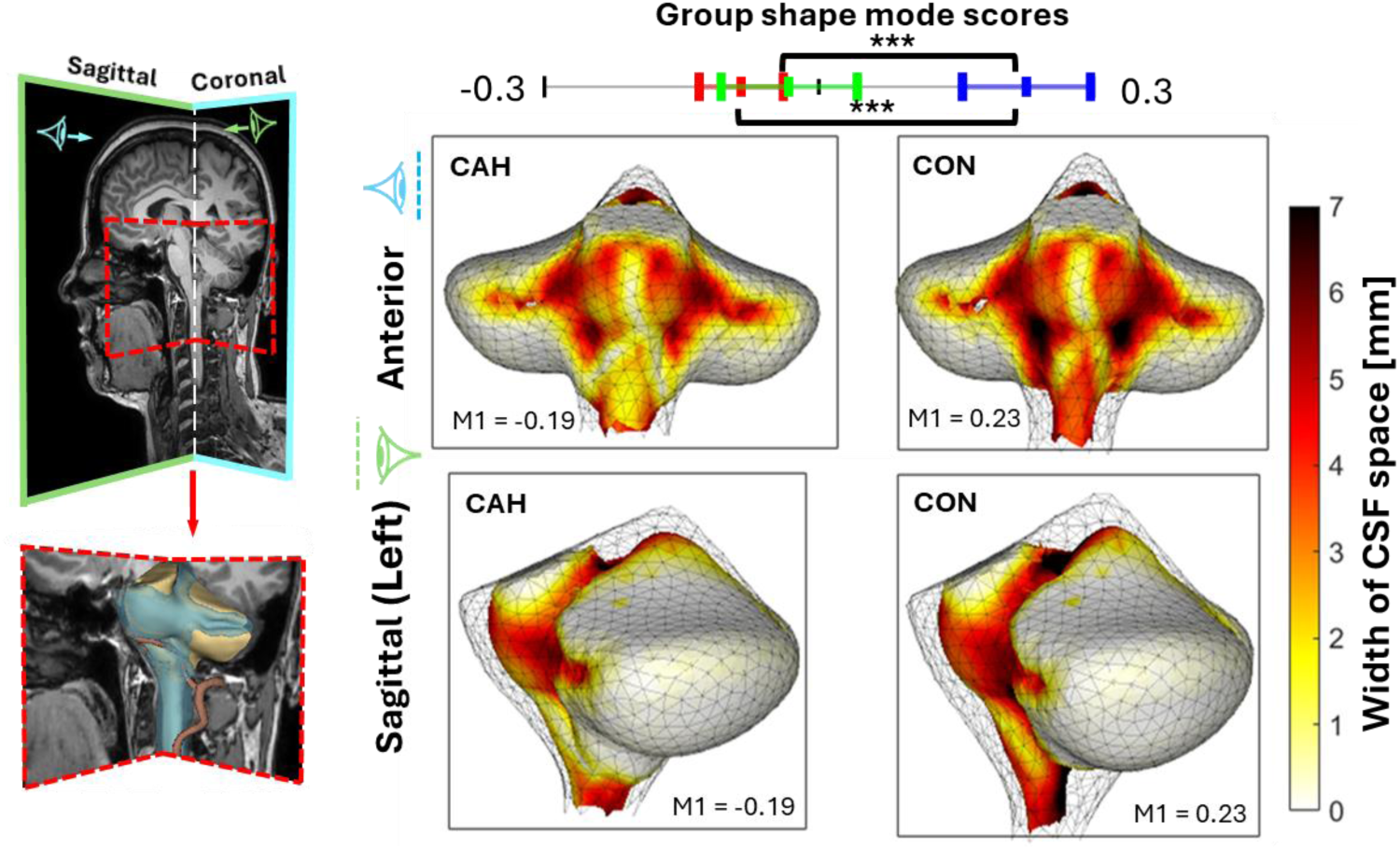
Heatmaps showing the difference in the width of the CSF space explained by the first shape mode score ±0.3 from the mean hindbrain shape. The anatomical images on the left indicate the directions the shape models are viewed, with the blue eye matching the anterior view, and the green eye the sagittal view. The line plot at the top shows the group average shape mode scores (short coloured vertical bars) ±95% CI (longer vertical bars). The plots contrast the shapes of the average control participant (CON), and Chiari patient with cough associated headache (CAH). Colour bar indicates the perpendicular width of the CSF space between the hindbrain, the vasculature, and boundaries of the posterior fossa (black wireframe mesh). Areas with no CSF space are white, the shading progresses from yellow, red, then black as the CSF space becomes greater than 7mm wide. Significant differences are indicated by the dashed arrows (*p<0.05, **p<0.01, and ***p<0.001).

**Figure 4.**
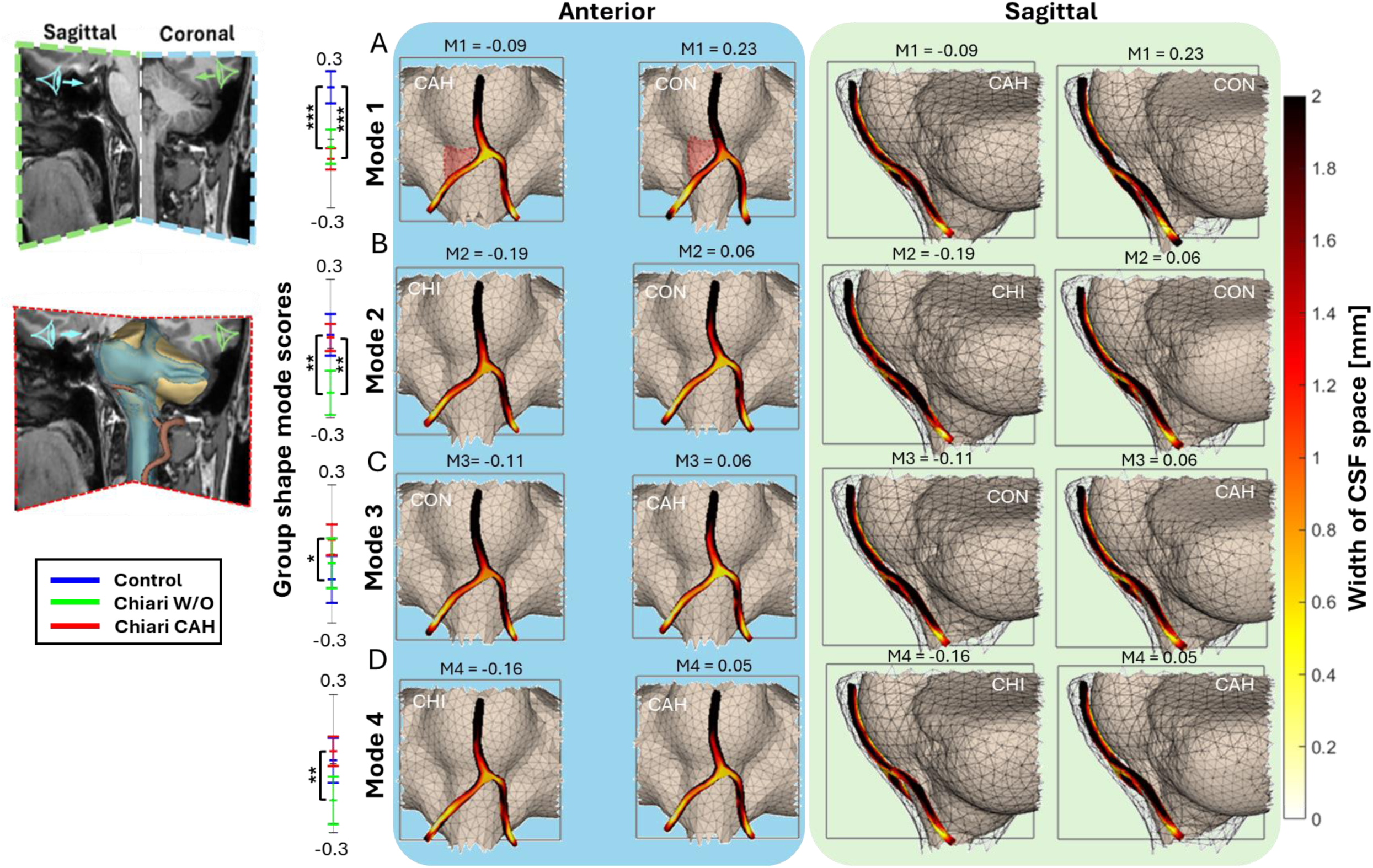
Heatmaps showing the difference in the width of the CSF space about the vertebrobasilar arteries that is explained by the first four shape mode scores (A-D). The line plot shows the group average shape mode scores ±95% CI. Plots A-D contrast the shapes of the average control participant (CON), or Chiari patient with (CAH) or without (CHI) cough-associated headache. The anatomical key highlights the directions the shape models are viewed, with the blue eye matching the anterior view, and the green eye the sagittal view. Colour bar indicates the width of the CSF space between the vasculature, hindbrain (beige mesh) and boundaries of the posterior fossa (black wireframe mesh). Areas with no CSF space are coloured white, the shading progresses from yellow, red, then black as the CSF space becomes greater than 2mm wide. Significant differences are indicated by the brackets (*p<0.05, **p<0.01, and ***p<0.001). The red patch highlighted on the medulla oblongata in row A indicates the general location of the right cranial nerves VI-X, which are symmetrical.

The same four shape modes also characterised the observed variation in the vertebrobasilar arteries (Figure 4). These four modes described changes in the location where the vertebral arteries converge to form the basilar artery, from midline to the lateral boundary between the pons and medulla in addition to the space available for CSF to flow around the arteries. Modes 1 & 2 were lower in patients without cough headache than controls (Figure 4A&B; Table 1). Mode 1 was lower, and Mode 3 was greater in patients with cough headache than controls (Figure 4A&C; Tabel 1). Greater scores for Modes 2 and 4 were found in patients with cough headache than without (Figure 4B&D, Table 1).

**Table 1.** Summary of group-wise comparison of the shape mode scores of healthy controls, patients with (CAH) and without (CHI) cough-associated headache. The group differences were calculated as the comparison group minus the reference group. Results are displayed as the mean difference ± 95% CI, and the effect size (Cohen’s d). Significant differences in shape mode scores are in bold and italicised.

| Ref. | Comparison | Shape Mode | Mean$\pm 95\%$ CI | Effect size [Cohen's d] | p-value |
| --- | --- | --- | --- | --- | --- |
| Control | CAH | *P1* | ***-0.31$\pm 0.10$*** | ***-3.01*** | ***<0.0001*** |
| | | P2 | -0.01$\pm 0.13$ | -0.09 | 1.00 |
|  |  | *P3* | ***0.17$\pm 0.15$*** | ***1.15*** | ***0.02*** |
| | | P4 | 0.04$\pm 0.14$ | 0.27 | 1.00 |
|  | CHI | *P1* | ***-0.26$\pm 0.13$*** | ***-2.51*** | ***<0.0001*** |
|  |  | *P2* | ***-0.25$\pm 0.16$*** | ***-1.89*** | ***0.001*** |
| | | P3 | 0.07$\pm 0.18$ | 0.47 | 1.00 |
| | | P4 | -0.17$\pm 0.18$ | -1.21 | 0.054 |
| CHI | CAH | P1 | -0.52$\pm 0.11$ | -0.50 | 0.70 |
|  |  | *P2* | ***0.24$\pm 0.14$*** | ***1.80*** | ***<0.001*** |
| | | P3 | 0.10$\pm 0.16$ | 0.68 | 0.33 |
|  |  | *P4* | ***0.21$\pm 0.15$*** | ***1.48*** | ***0.003*** |

### The effect of coughs on CSF flow

All CSF flow traces showed a similar pattern in response to coughs. Before the cough normal cardiac pulsations were present, with caudal flow during systole that transitions to cranial during diastole (Figure 5). During the initial phase of the cough cranial flow increased, and after the expiratory effort of the cough, CSF returned caudally. Immediately after the cough normal cardiac pulsations resumed. Figure 5 provides example CSF flow and velocity traces from a control and Chiari patient (with cough headache), measured at the foramen magnum. In these two subjects there were no differences in CSF *flow* before, during or after the cough. However, the CSF *velocities* were more than two times greater in the Chiari patient with cough headache than the control (Figure 5B).

**Figure 5.**
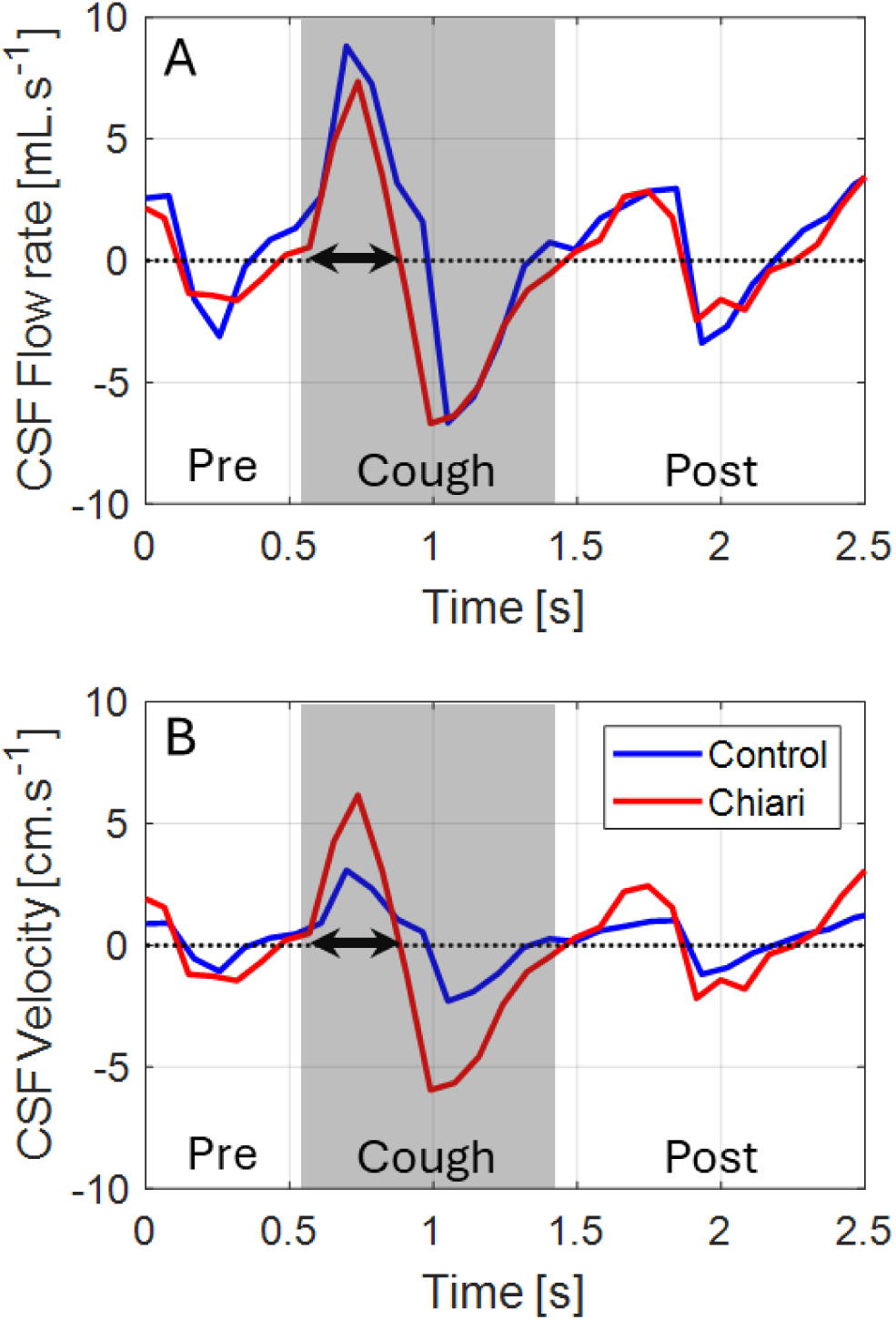
Example data showing of the effect of coughs on CSF flow (A) and average CSF velocity (B) taken from representative control and Chiari participants, at the level of the foramen magnum. The grey shading highlights the duration of a single cough. The black arrow highlights the duration of the expiratory effort of the cough in the Chiari participant.

In all locations both CSF flow and velocities were greater during coughs than before or after the cough (Figure 6). Below the obstruction at C3 both patients with and without cough headache had faster cranial CSF velocities than controls during coughs (Figure 6C; LMM; CAH, d = 1.23, p < 0.0001, no CAH, d = 3.42, p < 0.0001). Patients without cough headache also had greater cranial CSF flow than patients with cough headache, and controls, during coughs (Figure 6D; LMM; control, d = 2.31, p < 0.0001, CAH, d = 2.21, p < 0.0001).

**Figure 6.**
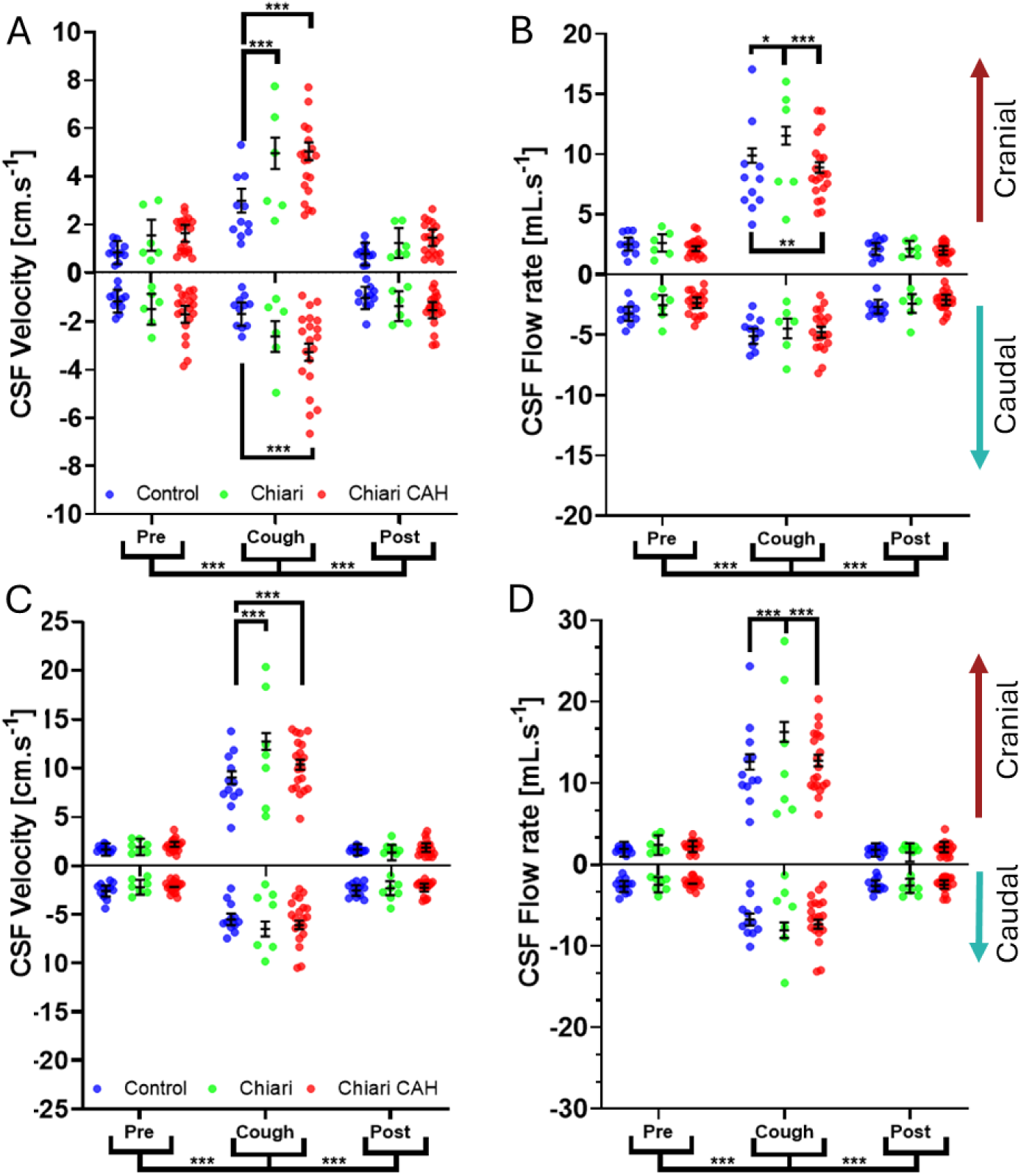
Group data showing the effect of cough on CSF dynamics at the foramen magnum (A,B), and at mid C3 (C,D). Panels A & C show the average CSF velocities, and the flow rates are displayed in B & D. Cranial CSF flow is positive (red arrows), and caudal flow is negative (blue arrows). The group data shows a point for the average response of each subject. Marginal means and 95% confidence intervals are estimated by the LMM. Significant differences are indicated by the brackets (*p<0.05, **p<0.01, and ***p<0.001).

At the foramen magnum, patients with Chiari had similar flow rates to healthy controls before and after coughs (Figure 6B). During coughs patients without cough headache had greater cranial flow than those with cough headache and controls (LMM; control, d = 1.64, p < 0.0001, CAH, d = 2.64, p = 0.003), and cranial flow was greater in controls than patients with cough headache (LMM; CAH, d = 1.00, p = 0.027). There were no differences in the CSF velocities before or after coughs (Figure 6A), however during coughs both patients with (LMM; cranial d = 2.46, p < 0.0001; caudal d = -1.95, p <0.0001) and without (LMM; cranial d = 2.37, p < 0.0001) cough headache had greater CSF velocities than healthy controls. Although, caudal CSF velocities of patients without cough headache and controls were similar (LMM; d = -1.14, p= 0.071).

### The effect of coughs on blood flow

Systolic flow in the vertebral arteries was lower in patients with cough headaches than healthy controls (LMM; d = -1.10, p = 0.014). There were no differences in the arterial flow rates between respiratory phases (Figure 7). On average coughs resulted in greater venous return during a cough than before (LMM; d = -0.26, p < 0.0001) or after the cough (LMM; d = -0.33, p < 0.0001). Additionally, venous return was greater in patients without cough headache than in patients with cough headache (LMM; d = 1.08, p = 0.042). There was an interaction effect showing that during a cough, patients without cough headache had greater venous return than both patients with cough headache (LMM; d = -1.69, p < 0.0001) and healthy controls (LMM; d = -1.25, p = 0.031).

**Figure 7.**
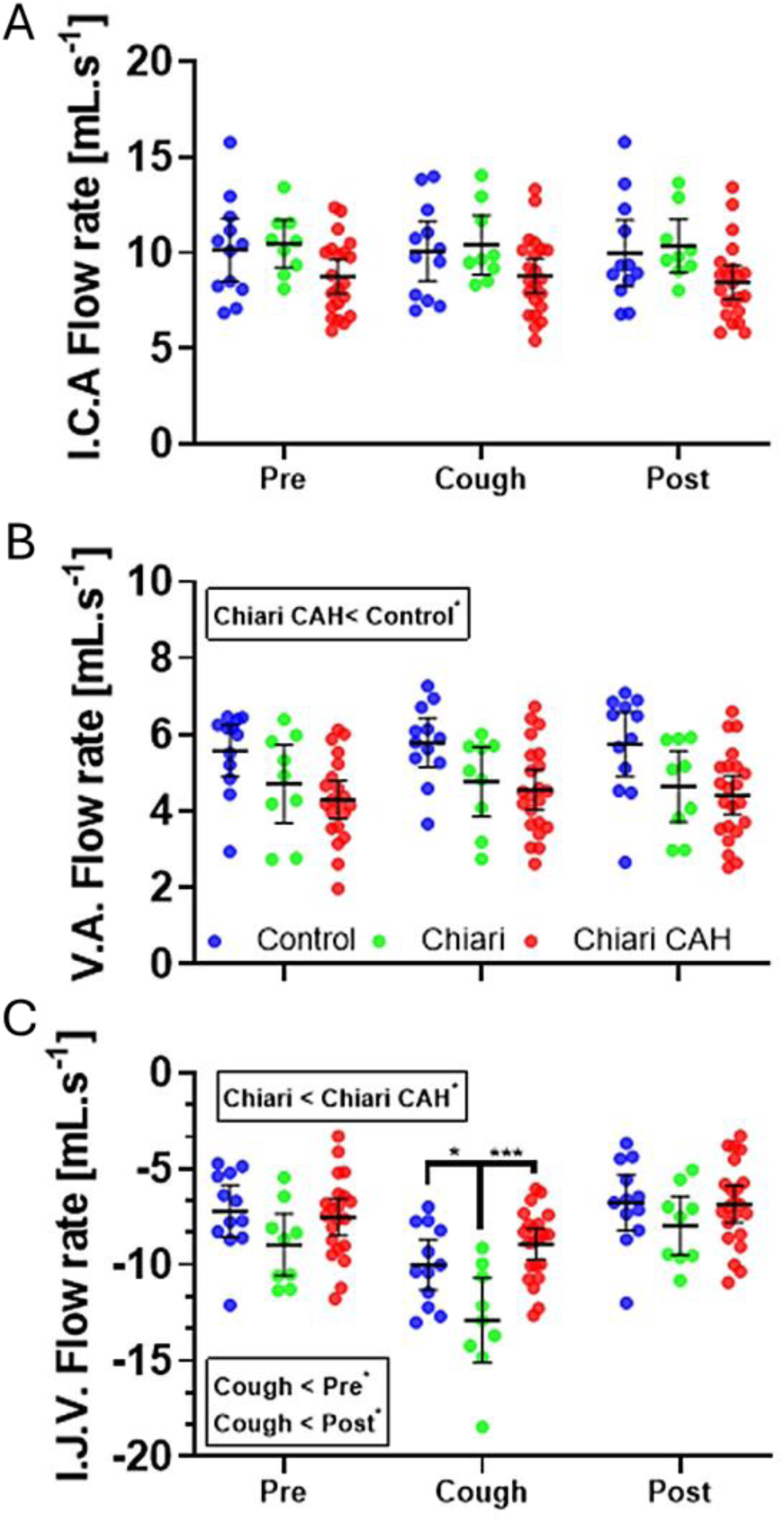
Group data showing the peak flow rate in the vertebral arteries (A), internal carotid arteries (ICA) (B), and the internal jugular veins (IJV) (C). Arterial flow into the cranium is signed positively, and venous outflow is signed negatively. Marginal means and 95% confidence intervals are estimated by the LMM. Significant differences are indicated by the brackets (*p<0.05, **p<0.01, and ***p<0.001)

## Discussion

In this study, real-time PC-MRI was used to assess how Chiari alters CSF flow during coughs. Our results showed CSF flow in patients was comparable to healthy controls and was not blocked or restricted during or after a cough, but the CSF velocities in Chiari patients were significantly higher than in controls. This suggests that a blockage/reduction in CSF flow is not required to trigger headache in patients, providing convincing evidence that the currently accepted mechanisms proposed by Williams (1980) are, at best, incomplete. We also used anatomical imaging to identify morphological features that characterise controls and patients with different symptoms. For the first time, we identified structural differences between patients with and without cough headache, whereby patients with cough headache had a narrower anterior CSF space adjacent to their vertebrobasilar arteries. This reduced anterior CSF space could contribute to cough-triggered headache by creating local stress concentrations acting on the arteries or adjacent neural structures during periods of high velocity CSF flow produced during coughs.

In contrast to the recent findings of Bhadelia et al. (2024), we did not find CSF flow decreased after coughs (Figures 5&6). This can be attributed to our participants performing single gentle coughs rather than repetitive strong coughs, but importantly, these were still able to cause headache in participants. During these coughs there was no change in arterial flow (Figure 7), suggesting that our manoeuvres also did not significantly affect arterial pressures, and were thus unlikely to trigger the baroreflex that varies the heart rate to maintain arterial pressures. Maximal expiratory efforts have been shown to trigger the baroreflex 10 – 20 s after the manoeuvre (Wei and Harris, 1982; Wei et al., 1983), similar to Phase III-IV of a Valsalva manoeuvre (Legg Ditterline et al., 2016). As post-cough was defined as 5 – 10 s after the coughing, this cardiovascular response would likely influence the findings of Bhadelia et al. (2024), making it unclear whether the difference in CSF flow in patients with cough headache was due to a transient obstruction or the coupled effect of the cranial morphology (Figures 3 and 4) and the baroreflex. Our protocol avoids the influence of the baroreflex.

Venous return during all respiratory phases was greatest in patients without cough headache (Figure 7C). This is in agreement with the findings of Alperin et al. (2015) that patients without cough headache had less resistance to venous outflow. Under the Monro-Kellie hypothesis, venous blood is ejected from the cranium in response to an influx of spinal CSF and epidural venous blood into the cranium (Lloyd et al., 2020). In patients without cough headache this reduced resistance to venous outflow may help compensate for the influx of CSF, preventing coughs from triggering headache.

Like previous morphological studies, our 3D shape model showed that in patients with Chiari the cerebellar tonsils occupy the posterior CSF space and reduce the CSF space anterior to the pons and medulla (García et al., 2021; Nwotchouang et al., 2021; Thakar et al., 2021). However, our 3D analyses suggest that the commonly used midsagittal measures overestimate the severity of the obstruction - even in patients with severe herniation, the anterolateral spaces remained patent (Figure 3), which allowed for relatively normal volumes of CSF to flow across the foramen magnum (Figure 6B). Patients with cough-associated headaches had significantly less CSF space near the vertebral and basilar arteries (Figure 4), and the convergence of the vertebrobasilar arteries was skewed laterally toward the junction between the medulla and pons. This positioned one vertebral artery close to the cranial nerves and the other crossing the anterior CSF space. These anatomical differences would likely increase the stresses acting on the blood vessel (depicted schematically in Figure 8), since the narrower CSF spaces increase fluid velocities and thus the wall shear stresses acting on the outer vessel wall (Figure 8B). Moreover, fluid drag forces from flow perpendicular to the artery could stretch or bend the vessels (Figure 8C). This may explain how coughs trigger headache, as the application of external pressure to (Ray and Wolff, 1940) or occlusion of (Nichols 3rd et al., 1990) the vertebral and basilar arteries, has been shown to generate occipital headaches. Interestingly these investigations demonstrated that depending on the focal point of the applied pressure/occlusion the headache location varied from the occiput to retro-orbital. Patient specific variation in the vascular and fossa morphology could produce localised stress concentrations (Figure 4), which may present as non-occipital headaches in patients with Chiari (Alperin et al., 2015; Bezuidenhout et al., 2019). This remains to be confirmed in future studies.

**Figure 8.**
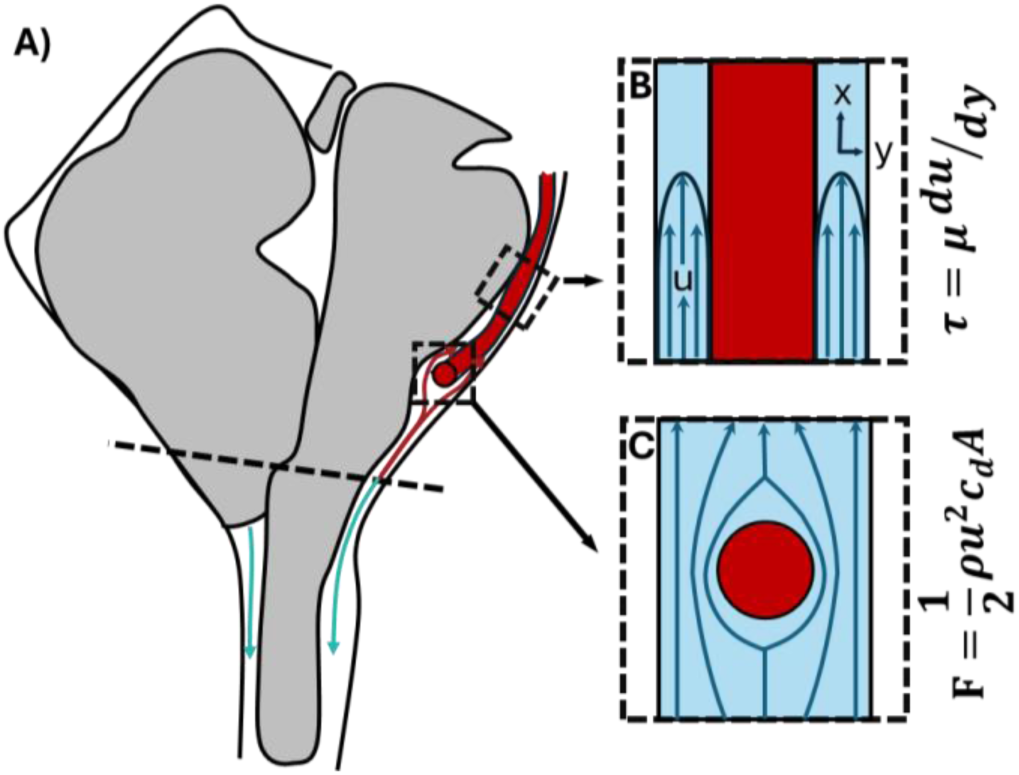
Schematic of hypothesised triggers of headache. A) In patients with Chiari the anterior CSF space stays patent, allowing for both cranial (red arrow) and caudal (blue arrow) flow during a cough. The narrow space between the hindbrain, arteries, and skull results in greater forces acting on the arteries, nerves, and meninges. Insets B and C provide examples of how patient anatomy may affect the forces acting on an artery. B) Both faster CSF velocities (u) and a narrower fluid channel (y) result in greater wall shear stresses (τ) on the surface of the artery due to viscous forces (µ = dynamic viscosity; ^*du*^_*dy*_ = velocity gradient). C) CSF drag forces (F) acting on blood vessels due to fluid inertia would be greatest (c_d_: 0.82-1.17) as it transitions from being in line with to perpendicular to the flow field (c_d_ = drag coefficient; A = cross-sectional area of the vessel perpendicular to the flow). These forces could displace or stretch the blood vessels or adjacent neural structures, contributing to headaches.

### Implications for Chiari treatment

Our study suggests two factors that contribute to patient symptoms. Firstly, patients have anterior CSF flow velocities twice as fast as in the healthy population. Secondly, that patients with cough-associated headache have a narrower CSF space around the vertebrobasilar arteries than patients without cough headaches. Both of these would increase forces on these vessels, which has been shown to generate occipital headache symptoms (Nichols 3rd et al., 1990; Ray and Wolff, 1940). If this mechanism is correct, it implies that for successful resolution of these headaches, surgical treatment needs to enlarge the anterior CSF space to reduce loading on these vessels. The most common treatment for Chiari is decompression surgery, which expands the posterior fossa and normalises CSF flow. It is not currently a focus of this surgery to specifically enlarge the anterior CSF space, and the extent to which this occurs may explain the variation in headache resolution after surgery. Testing this hypothesis requires more detailed analysis of CSF dynamics in the anterior CSF space before and after decompression surgery in people with cough-associated headache.

### Study Limitations

Due to the respiratory sensor used, the strength of participant coughs was not able to be quantified, so differences in expiratory effort may contribute to the variance in the magnitude of CSF flow/velocity reported (Lloyd et al., 2020). The CSF flow rates during coughs at mid-C3 where approximately half (Figure 6D; average cranial cough flow = 11±1mL.s^-1^), compared to our previous measurements in healthy controls (Lloyd et al. 21±5mL.s^-1^). As gentle coughs were performed in this study, our results may underestimate the effect of coughs on CSF velocities, and this should be considered when interpreting our data.

Although we have identified morphological differences between patients with (N=20) and without (N=7) cough-associated headache, our sampled population contains more participants with cough-associated headache than without. The large effect size of these comparisons provides confidence that these morphological characteristics genuinely reflect the patient groups (Figure 4). The small sample of patients without cough headache may bias the mean CSF flow rates and velocities, given the large spread of these data (Figures 6B, C and D), and a larger study should be conducted to demonstrate that these findings are representative of the broader Chiari patient population. Additionally, only a small sample of our cohort has undergone posterior fossa decompression, preventing the analysis of anterior vs posterior CSF flow changes and surgical outcome.

## Conclusion

This study demonstrated that Chiari does not markedly block CSF flow during or after a cough, with lateral flow compensating for mid-sagittal CSF space restriction by the cerebellar tonsils. However, the overcrowded anterior CSF space results in greater CSF velocities in patients than healthy controls. The shape analysis highlighted that those patients with cough-associated headaches had a narrower anterior CSF space, particularly around the vertebrobasilar arteries, than patients without cough headache. The high velocity CSF flow during coughs and the reduced CSF space may increase arterial stress and trigger the headaches. Further analysis of cranial morphology and CSF velocities associated with outcomes of decompression surgery on headache would help establish the validity of this hypothesised headache mechanism, and the utility of these imaging metrics for diagnosis and the prediction of surgical outcomes in Chiari patients.

## Abbreviations

(CSF): Cerebrospinal Fluid
(MRI): Magnetic Resonance Imaging
(PC-MRI): Phase-Contrast Magnetic Resonance Imaging
(IJV): Internal Jugular Veins
(LMM): Linear Mixed Model.

## Additional information

### Author contributions

R.A.L., L.E.B., M.A.S conceived and designed the experiments. R.A.L., and A.D.M., performed the experiments. All authors contributed to the interpretation of the data and writing the paper. All authors approved the final version of the manuscript submitted for publication and agree to be accountable for all aspects of the work. All persons designated as authors qualify for authorship, and all who qualify for authorship are listed.

### Ethics Statement

All experiments were conducted in accordance with the policies of Neuroscience Research Australia, and the University of New South Wales (UNSW). Experimental protocols were approved by the South Eastern Sydney Local Health District Human Research Ethics Committee (Ref No: 2019/ETH04885, Approval date: 31^st^ Aug 2016) and were conducted according to the Declaration of Helsinki (2013), except for clause 35. All participants gave written informed consent. Data of individual participants are not publicly available in compliance with the approved ethics.

### Data availability statement

The weights for the trained CSF PC-MRI, and posterior fossa nnU-net models are available at *[To be made available before peer review]*. The data that support the findings of this study are not publicly available due to restrictions of the approved ethics.

## Acknowledgements

For the purposes of open access, the author has applied a CC BY public copyright licence to any Author Accepted Manuscript version arising from this submission. The authors acknowledge the facilities and scientific and technical assistance of NeuRA Imaging, a node of the National Imaging Facility, a National Collaborative Research Infrastructure Strategy (NCRIS) capability.

## References

1. Abdul-Rahman, H., Gdeisat, M., Burton, D., Lalor, M., 2005. Fast Three-Dimensional Phase-Unwrapping Algorithm Based on Sorting by Reliability Following a Non-Continuous Path. SPIE.

2. Alperin, N., Loftus, J.R., Oliu, C.J., Bagci, A.M., Lee, S.H., Ertl-Wagner, B., Sekula, R., Lichtor, T., Green, B.A., 2015. Imaging-Based Features of Headaches in Chiari Malformation Type I. Neurosurgery 77, 96–103. 10.1227/NEU.0000000000000740

3. Bezuidenhout, A.F., Chang, Y.M., Heilman, C.B., Bhadelia, R.A., 2019. Headache in Chiari Malformation. Neuroimaging Clin N Am 29, 243–253. 10.1016/j.nic.2019.01.005

4. Bezuidenhout, A.F., Khatami, D., Heilman, C.B., Kasper, E.M., Patz, S., Madan, N., Zhao, Y., Bhadelia, R.A., 2018. Relationship between Cough-Associated Changes in CSF Flow and Disease Severity in Chiari I Malformation: An Exploratory Study Using Real-Time MRI. American Journal of Neuroradiology 39, 1267. 10.3174/ajnr.A5670

5. Bhadelia, R.A., Ibrahimy, A., Al Samman, M.M., Ebrahimzadeh, S.A., Zhao, Y., Loth, F., 2024. Transient Decrease in Cerebrospinal Fluid Motion Is Related to Cough-Associated Headache in Chiari I Malformation. World Neurosurg 189, e709–e717. 10.1016/j.wneu.2024.06.152

6. Bhadelia, R.A., Patz, S., Heilman, C., Khatami, D., Kasper, E., Zhao, Y., Madan, N., 2016. Cough-Associated Changes in CSF Flow in Chiari I Malformation Evaluated by Real-Time MRI. American journal of neuroradiology 37, 825–830. 10.3174/ajnr.A4629

7. Bydder, M., 2019. Unwrap [Computer Software]: Matlab C++ Mex Wrappers to the Phase Unwrapping Code at https://Github.Com/Geggo/Phase-Unwrap. https://github.com/marcsous/unwrap

8. Cates, J., Elhabian, S., Whitaker, R., 2017. Chapter 10 - Shapeworks: Particle-Based Shape Correspondence and Visualization Software, in: Zheng, G., Li, S., Székely, G. (Eds.), Statistical Shape and Deformation Analysis. Academic Press, pp. 257–298. 10.1016/B978-0-12-810493-4.00012-2

9. Dawes, B.H., Lloyd, R.A., Rogers, J.M., Magnussen, J.S., Bilston, L.E., Stoodley, M.A., 2019. Cerebellar Tissue Strain in Chiari Malformation with Headache. World Neurosurg 130, e74–e81. 10.1016/j.wneu.2019.05.211

10. Du Boulay, G., O’Connell, J., Currie, J., Bostick, T., Verity, P., 1972. Further Investigations on Pulsatile Movements in the Cerebrospinal Fluid Pathways. Acta Radiol Diagn (Stockh) 13, 496–523.

11. Fedorov, A., Beichel, R., Kalpathy-Cramer, J., Finet, J., Fillion-Robin, J.-C., Pujol, S., Bauer, C., Jennings, D., Fennessy, F., Sonka, M., Buatti, J., Aylward, S., Miller, J.V., Pieper, S., Kikinis, R., 2012. 3D Slicer as an Image Computing Platform for the Quantitative Imaging Network. Magnetic resonance imaging 30, 1323–1341. 10.1016/j.mri.2012.05.001

12. García, M., Eppelheimer, M.S., Houston, J.R., Houston, M.L., Nwotchouang, B.S.T., Kaut, K.P., Labuda, R., Bapuraj, J.R., Maleki, J., Klinge, P.M., Vorster, S., Luciano, M.G., Loth, F., Allen, P.A., 2021. Adult Age Differences in Self-Reported Pain and Anterior CSF Space in Chiari Malformation. The Cerebellum. 10.1007/s12311-021-01289-w

13. Hersh, D.S., Groves, M.L., Boop, F.A., 2019. Management of Chiari Malformations: Opinions from Different Centers—a Review. Childs Nerv Syst 35, 1869–1873.

14. Isensee, F., Jaeger, P.F., Kohl, S.A.A., Petersen, J., Maier-Hein, K.H., 2021. Nnu-Net: A Self-Configuring Method for Deep Learning-Based Biomedical Image Segmentation. Nat Methods 18, 203–211. 10.1038/s41592-020-01008-z

15. Legg Ditterline, B.E., Aslan, S.C., Randall, D.C., Harkema, S.J., Ovechkin, A.V., 2016. Baroreceptor Reflex During Forced Expiratory Maneuvers in Individuals with Chronic Spinal Cord Injury. Respiratory Physiology & Neurobiology 229, 65–70. 10.1016/j.resp.2016.04.006

16. Leung, V., Magnussen, J.S., Stoodley, M.A., Bilston, L.E., 2016. Cerebellar and Hindbrain Motion in Chiari Malformation with and without Syringomyelia. J Neurosurg Spine 24, 546–555. 10.3171/2015.8.spine15325

17. Lloyd, R.A., Butler, J.E., Gandevia, S.C., Ball, I.K., Toson, B., Stoodley, M.A., Bilston, L.E., 2020. Respiratory Cerebrospinal Fluid Flow Is Driven by the Thoracic and Lumbar Spinal Pressures. J Physiol 598, 5789–5805. 10.1113/JP279458

18. Meadows, J., Kraut, M., Guarnieri, M., Haroun, R.I., Carson, B.S., 2000. Asymptomatic Chiari Type I Malformations Identified on Magnetic Resonance Imaging. J Neurosurg 92, 920–926. 10.3171/jns.2000.92.6.0920

19. Milhorat, T.H., Chou, M.W., Trinidad, E.M., Kula, R.W., Mandell, M., Wolpert, C., Speer, M.C., 1999. Chiari I Malformation Redefined: Clinical and Radiographic Findings for 364 Symptomatic Patients. Neurosurgery 44. 10.1097/00006123-199905000-00042

20. Nichols 3rd, F., Mawad, M., Mohr, J., Stein, B., Hilal, S., Michelsen, W.J., 1990. Focal Headache During Balloon Inflation in the Internal Carotid and Middle Cerebral Arteries. Stroke 21, 555–559.

21. Nwotchouang, B.S.T., Ibrahimy, A., Loth, D.M., Labuda, E., Labuda, N., Eppleheimer, M., Labuda, R., Bapuraj, J.R., Allen, P.A., Klinge, P., Loth, F., 2021. Imaging and Health Metrics in Incidental Cerebellar Tonsillar Ectopia: Findings from the Adolescent Brain Cognitive Development Study (Abcd). Neuroradiology 63, 1913–1924. 10.1007/s00234-021-02759-y

22. Parker, S.L., Godil, S.S., Zuckerman, S.L., Mendenhall, S.K., Wells, J.A., Shau, D.N., McGirt, M.J., 2013. Comprehensive Assessment of 1-Year Outcomes and Determination of Minimum Clinically Important Difference in Pain, Disability, and Quality of Life after Suboccipital Decompression for Chiari Malformation I in Adults. Neurosurgery 73, 569–581. 10.1227/neu.0000000000000032

23. Ray, B.S., Wolff, H.G., 1940. Experimental Studies on Headache: Pain-Sensitive Structures of the Head and Their Significance in Headache. Archives of Surgery 41, 813–856.

24. SCI Institute, 2021. Shapeworks (Version 6.1) [Computer Software]. GitHub https://github.com/SCIInstitute/ShapeWorks/releases/tag/v6.1.0

25. Sharpey-Schafer, E., 1953. Effects of Coughing on Intra-Thoracic Pressure, Arterial Pressure and Peripheral Blood Flow. J Physiol 122, 351.

26. Thakar, S., Kanneganti, V., Talla Nwotchouang, B.S., Salem, S.J., Eppelheimer, M., Loth, F., Allen, P.A., Aryan, S., Hegde, A.S., 2021. Are Two-Dimensional Morphometric Measures Reflective of Disease Severity in Adult Chiari I Malformation? World Neurosurg. 10.1016/j.wneu.2021.10.138

27. Usubiaga, J.E., Moya, F., Usubiaga, L.E., 1967. Effect of Thoracic and Abdominal Pressure Changes on the Epidural Space Pressure. BJA 39, 612–618. 10.1093/bja/39.8.612

28. Wang, B., Wang, C., Zhang, Y.-W., Liang, Y.-C., Liu, W.-H., Yang, J., Xu, Y.-L., Wang, Y.-Z., Jia, W.-Q., 2023. Long-Term Outcomes of Foramen Magnum Decompression with Duraplasty for Chiari Malformation Type I in Adults: A Series of 297 Patients. Neurosurg Focus 54, E5. 10.3171/2022.12.FOCUS22627

29. Wei, J.Y., Harris, W.S., 1982. Heart Rate Response to Cough. J Appl Physiol 53, 1039–1043.

30. Wei, J.Y., Rowe, J.W., Kestenbaum, A.D., Ben-Haim, S., 1983. Post-Cough Heart Rate Response: Influence of Age, Sex, and Basal Blood Pressure. American Journal of Physiology-Regulatory, Integrative and Comparative Physiology 245, R18–R24.

31. Williams, B., 1980. Cough Headache Due to Craniospinal Pressure Dissociation. Archives of Neurology 37, 226–230. 10.1001/archneur.1980.00500530064010

32. Zhou, L.-N., Xiao, X., Chen, X.-Y., Gu, S.-X., Liu, X.-D., Shou, J.-J., Gu, W.-T., Che, X.-M., Zhao, J.-L., Xie, R., 2024. The Surgical Strategy Cerebrospinal Fluid Decompression Facilitates Outcomes of Adults with Chiari Malformation Type I: An Observational, Real-World, Single-Center Study of 528 Patients. World Neurosurg 189, e841–e856. 10.1016/j.wneu.2024.07.016

