## Supplemental Figures S1-3 for "Chiari malformation does not obstruct cerebrospinal fluid flow during coughs"

### Methods

#### Training data

The model for segmenting the posterior cranial fossa regions of interest (ROI; CSF, soft tissue, and vertebrobasilar arteries) were trained from the manual segmentation of 13 subjects, and validated against the ROI of 3 withheld subjects. The foramen magnum real-time phase contrast (PC) MRI ROI was trained on 46 scans and validated against 11 withheld data sets. The mid-C3 PC-MRI ROI used 36 scans and was validated with 10 withheld scans.

#### Training scheme

To automate segmentation of the posterior cranial fossa, and real-time PC- MRI CSF flow, the nnU-Net framework (Isensee et al., 2021) was used to configure a 3D U-Net (Çiçek et al., 2016) CNN. Training was performed on a NVIDIA RTX3080 GPU with 10 GB memory. For the anatomical models 3D-isotropic T1-weighted images were used as a single channel for training the network. Training was conducted on 150 epochs of 250 minibatches. For PC-MRI taken at the foramen magnum (FM) and mid-C3, the magnitude, phase-magnitude, and phase images were used as individual contrast models during training. The PC-MRI data was restructured to have time frames in the 3$^{rd}$ dimension, of the image volumes. Training was conducted on 1000 epochs of 250 minibatches. nnU-Net applied data augmentation (image rotation/scaling, additive noise, and contrast enhancements) during training to reduce overfitting. For a detailed overview of the nnU-Net framework, see Isensee et al. (2021).

The Dice coefficient (DSC; Eqn. 1), average symmetric surface distance (ASSD; Eqn. 2), maximum symmetric surface distance (MSSD; Eqn. 3), and volume error, ($V_{Error}$; Eqn. 4) were used to assess the similarity of each models predicted and reference ROI. The DSC quantifies the overlap of voxels between the ground truth (G) and predicted (P) labels, with 1 indicating perfect agreement. ASSD and MSSD describes the average and maximum Euclidian distance (d) of voxels (p) on the surface of the predicted ($P_b$), to the corresponding voxels (g) on the boundary of the reference label ($G_b$), respectively. $V_{Error}$ measures the absolute difference in the volume of the predicted and reference labels, where N is the number of voxels, and $V_{res}$ is the volume of a voxel.

$$DSC = \frac{2|G \cap P|}{|G|+|P|} \quad \text{Eqn. 1}$$

$$ASSD = \frac{\sum d(p, G_b) + \sum d(g, P_b)}{|G_b|+|P_b|} \quad \text{Eqn. 2}$$

$$MSSD = \max\left[\max[d(p, G_b)], \max[d(g, P_b)]\right] \quad \text{Eqn. 3}$$

$$V_{Error} = |N_G - N_p| * V_{res} \quad \text{Eqn. 4}$$

### Results

#### Anatomical segmentation

**Table S1. Summary statistics of agreement between manual segmentation and predicted labels. Results shown as mean ± S.D.**

| ROI | DICE | ASSD [mm] | MSSD [mm] | Volume error [cm$^3$] |
|---|---|---|---|---|
| CSF | 0.88±0.05 | 1.0±1.0 | 32.0±2 | 1.0±1.0 |
| Soft tissue | 0.98±0.01 | 0.5±0.3 | 20.0±10.0 | 0.4±3 |
| Vertebrobasilar | 0.60±0.30 | 3.0±4.0 | 23.0±14.0 | 0.5±0.6 |

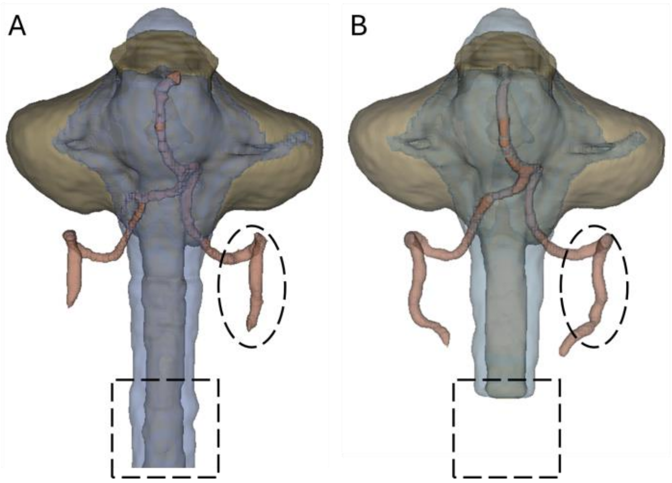


**Figure S1. Example of the ground truth (A) and predicted label (B) from validation set with the lowest Dice score. The volumes of the CSF (blue), soft tissues (brown), and arteries (red) are in agreement. The lower dice scores can be attributed to the manual segmentation extending to mid-C5 (dashed box) in the ground truth data (A), but only mid-C3 in the predicted labels (B). Additionally, outside of the CSF there was less contrast between the arteries and surrounding tissues, leading to errors in the prediction of the vertebral arteries (dashed circle). However, the branches outside of the CSF are excluded from analysis in this study and will not bias our results.**

### CSF flow segmentation

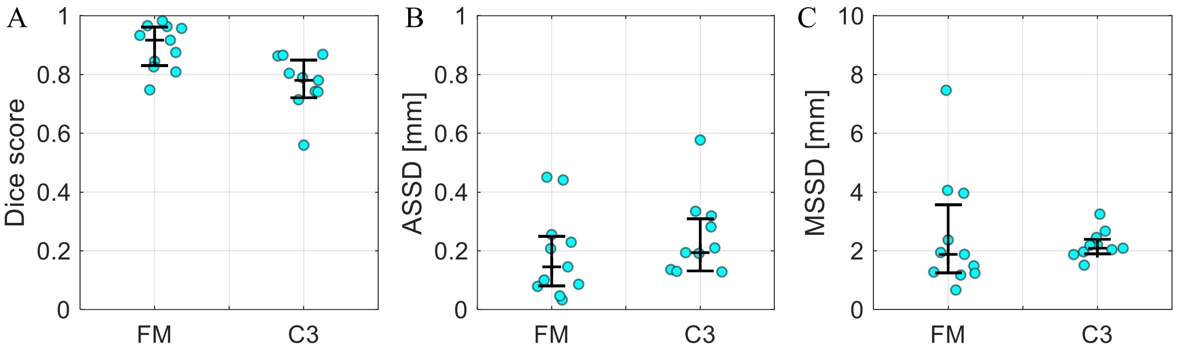


**Figure S2. Summary statistics (median and IQR) of U-Net segmentation of CSF flow at the level of the foramen magnum (FM) and Mid C3. A) shows the Dice score, B) show the average symmetric surface distance (ASSD), and C) the maximum symmetric surface distance.**

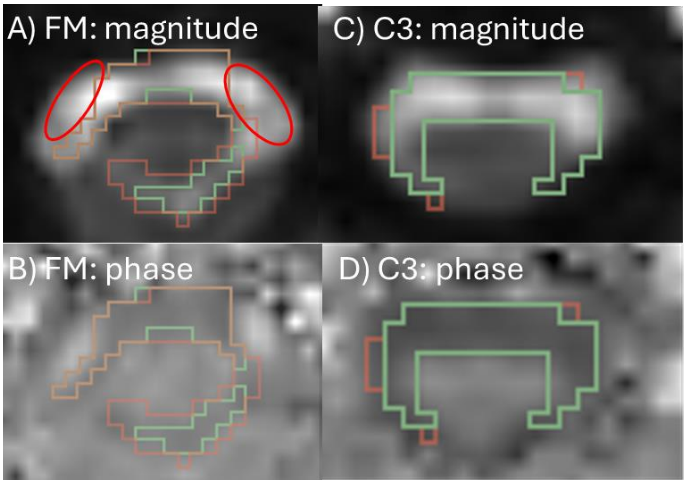


**Figure S3. Example predicted (green) and ground truth (orange) CSF segmentations of PC-MRI taken at the level of the foramen magnum (FM, A&B), and at mid-C3 (C&D). At the foramen magnum the predicted mask correctly excluded the vertebral arteries (red ellipses), however the model would underpredict the CSF area in narrow CSF spaces (B) with a low magnitude signal (A). At mid-C3 the largest differences in CSF masks were when the ground truth masks underestimated the CSF area where there was low velocities and poor contrast.**

### References

Çiçek, Ö., Abdulkadir, A., Lienkamp, S.S., Brox, T., Ronneberger, O., 2016 3D U-Net: Learning Dense Volumetric Segmentation from Sparse Annotation. In International conference on medical image computing and computer-assisted intervention.

Isensee, F., Jaeger, P.F., Kohl, S.A., Petersen, J., Maier-Hein, K.H., 2021. Nnu-Net: A Self-Configuring Method for Deep Learning-Based Biomedical Image Segmentation. Nature methods 18, 203-211.

#### Funding

Rob Lloyd and this research were supported by a National Health and Medical Research Council (NHMRC) Ideas grant (APP2011940). Adam Martinac was also supported by the NHMRC Ideas grant (APP2011940).

#### Acknowledgements

For the purposes of open access, the author has applied a CC BY public copyright licence to any Author Accepted Manuscript version arising from this submission. The authors acknowledge the facilities and scientific and technical assistance of NeuRA Imaging, a node of the National Imaging Facility, a National Collaborative Research Infrastructure Strategy (NCRIS) capability.
